# Phenotypic and Genetic Associations Between Cardiovascular Disease Subtypes and Alzheimer’s Disease

**DOI:** 10.1101/2025.08.29.25334750

**Authors:** Aili Toyli, Chen Zhao, Kuan-Jui Su, Hui Shen, Hong-Wen Deng, Qing-Hui Chen, Qiuying Sha, Weihua Zhou

**Author notes:** Corresponding authors: Weihua Zhou, Ph.D., Department of Applied Computing, Michigan Technological University, 1400 Townsend Dr, Houghton, MI, 49931, USA.

## Abstract

**Background:** Cardiovascular disease (CVD) and Alzheimer’s disease (AD) are major public health concerns that share overlapping risk factors and potential mechanistic pathways. While vascular contributions to cognitive decline are well-documented, the specific relationships between AD and different CVD subtypes remain poorly understood.

**Methods:** We examined associations between AD and 11 CVD subtypes using logistic regression models in two large biobanks: the UK Biobank (n = 502,133) and the All of Us Research Program (n = 287,011). Models were adjusted for demographic, lifestyle, and clinical covariates. We also explored genetic overlap between AD and CVD traits through colocalization of significant single nucleotide polymorphisms (SNPs) (p < 5×10^−8^) using genome-wide association study (GWAS) data.

**Results:** Most CVD subtypes were significantly associated with AD in both cohorts. Hypotension had the strongest and most consistent association, followed by hypertension and cerebral infarction. Acute myocardial infarction was the only subtype not significantly linked to AD. Genetic analyses revealed shared loci between AD and CVD-related traits, particularly in regions near *APOE, MAPT*, and genes influencing myocardial structure and vascular function.

**Conclusions:** This study identifies subtype-specific CVD associations with AD across two diverse cohorts and highlights shared genetic architecture underlying heart–brain interactions. These findings underscore the importance of vascular health in AD risk and suggest that certain CVD subtypes, especially hypotension, may play underrecognized roles in cognitive decline.

## Introduction

A growing body of evidence supports a strong and bidirectional relationship between cardiovascular and brain health^1^. The brain relies heavily on the heart, receiving 15% of the body’s cardiac output and 20% of its oxygen supply^2^, while the heart’s function is regulated by the nervous system^3^. Among the many intersections between these two systems, the link between cardiovascular disease (CVD) and Alzheimer’s disease (AD)—the most common form of dementia—is of particular interest^1,2^.

Both CVD and AD are age-associated chronic conditions that share overlapping risk factors including obesity, diabetes, smoking, environment, stress and sex differences^4,5^. Pathophysiologically, vascular damage from conditions like hypertension can disrupt the blood-brain barrier and lead to cerebral small vessel disease, manifesting as features such as white matter hyperintensities and microinfarcts^6^, which are associated with cognitive decline^7^. Conversely, forms of AD pathology like β-amyloid (Aβ) plaques and tau deposition may impair the regulation of autonomic nervous system and induce damage to cardiovascular function^4,8^. Despite this known interplay, the differential impact of specific CVD subtypes on AD risk remains poorly understood.

Large-scale population biobanks like the UK Biobank (UKB)^9^ and the All of Us Research Program (AoU)^10^ offer an unprecedented opportunity to clarify these relationships through robust, high-powered analyses across diverse populations^11^. Prior studies have examine individual CVD traits in relation to cognitive decline or AD, but comprehensive, subtype-level comparisons are limited^12–16^. This study aims to fill that gap by examining associations between AD and a wide range of CVD subtypes in both UKB and AoU, while also exploring shared genetic architecture through genome-wide association study (GWAS) data. Understanding which CVD subtypes are most strongly linked to AD may reveal key mechanistic pathways and inform targeted prevention strategies.

## Methods

### Study Population

We utilized data from two large, population-based biobanks: the UK Biobank (UKB) and the All of Us Research Program (AoU). UKB is a prospective cohort of 502,133 participants recruited from 22 centers across the UK between 2006 and 2010. The UKB resource is open to all bona fide researchers. Full details of its design and conduct are available online (https://www.ukbiobank.ac.uk). UKB received ethical approval from the National Health Service (NHS) Research Ethics Service (11/NW/0382); we conducted this analysis under application number 61915. All participants provided written informed consent, and the research was conducted in line with the Declaration of Helsinki.

AoU is a U.S.-based cohort established in 2015, with over 746,000 enrolled participants, of whom 287,011 had linked electronic health record (EHR) and survey data necessary for this study. All of Us Research Program is reviewed by an internal human subjects review board and all participants provide informed consent. Secondary analyses of de-identified data from AoU is not considered human subjects research.

### Phenotype Ascertainment

AD and CVD subtypes were identified via International Classification of Diseases, 10th Revision (ICD-10)^17^ codes from EHRs. We included the 11 CVD subtypes with ≥10,000 cases in UKB: hypertension (I10), hypotension (I95), angina pectoris (I20), acute myocardial infarction (I21), pulmonary embolism (I26), atrial fibrillation (I48), heart failure (I50), atrioventricular/left bundle-branch blockages, which are subsequently referred to as “blockage” (I44), chronic rheumatic heart disease (I05– I09), chronic ischemic heart disease (I25), and cerebral infarction (I63). AD was defined using ICD-10 code G30.

### Statistical Analysis

We performed cross-sectional logistic regression to estimate the odds ratios (ORs) of AD associated with each CVD subtype. Models were adjusted for key covariates affecting heart and brain health: age at assessment, sex, smoking status, education (age completed full-time education), depression status, physical activity (moderate and vigorous days/week), alcohol consumption, ethnicity, body mass index (BMI), annual income, and type 2 diabetes status^1,4,18,19^.

In UKB, most covariates were self-reported via baseline touchscreen questionnaire at the first visit to the assessment center^20^. BMI was calculated from height and weight measurements at first visit, diabetes status was determined through clinical records (E11), and depression was classified as “Yes” for a positive response to any type of depression or bipolar disorder, and “No” otherwise. All ethnic subgroups within the “White”, “Black”, “Mixed”, and “Asian” categories were described as their broader classification. All other variables retained their original levels from the UK BioBank^21^. Missing continuous covariate values were imputed using the median^22^ and categorical NAs were grouped as “Prefer not to answer.”

The same models were applied using AoU data where possible. We were not able to calculate the OR for pulmonary embolism, as there were only two positive cases with AD, making estimates highly unstable. Due to limited data availability, physical activity was excluded. BMI was averaged from all values reported for each participant. Age was estimated as years since birth as of 2025. Depression and diabetes were identified using ICD-10 codes. Ethnicity, smoking, income, alcohol use, and sex at birth were derived from self-reports, with “Prefer not to answer” and “Skip” responses combined. All individuals who listed multiple ethnicities were classified as “Mixed”.

### Genetic Analysis

To explore shared genetic architecture, we examined proximity between significant single nucleotide polymorphisms (SNPs) from UKB CVD GWASs performed by Backman et al.^23^ and SNPs from published GWASs related to brain structure and function included in the National Human Genome Research Institute-European Bioinformatics Institute (NHGRI-EBI) Catalog^24^ under the topics of dementia and all child traits, including AD, psychological disorder traits, and traits related to amyloid and tau levels. We assessed the reverse relationship too, as AD GWAS results from UKB were compared to catalog SNPs and cardiovascular disease traits, traits related to diastolic and systolic blood pressure, electrocardiogram derived traits, heart function, cardiac MRI values, heart rate, and heart shape measurements.

We also compared AD GWAS results from the catalog to GWAS results for 82 cardiac magnetic resonance imaging (CMR) traits in UKB. These CMR traits were derived through previous research and were return to UKB in the category “Cardiac and aortic function #1”. The cohort used for GWAS analysis for these traits was filtered to exclude participants with varying genetic and reported sex, non-white British ancestry, sex chromosome aneuploidy, and relatives. After filtering, 26,335 participants remained in the discovery dataset. We adjusted for the same covariates considered in previous research by Zhao et al^25^.

All GWASs used GRCh38 genome build^26^. We filtered for SNPs with p < 5×10⁻⁸ and identified colocalizations as SNP pairs located within 50 kb on the same chromosome. We then utilized NHGRI-EBI to search for genes containing these SNPs.

## Results

### Demographic Characteristics

In both the UK Biobank (UKB) and All of Us (AoU) cohorts, participants with Alzheimer’s disease (AD) were generally older, more likely to have diabetes, and had lower educational attainment and income compared to those without AD. AD cases were also more likely to be former or current smokers and less likely to report frequent alcohol consumption. BMI differences were minimal between groups. Ethnic representation was broader in AoU, while UKB was predominantly White (94.1%). These findings are summarized in Table 1 and Table 2.

**Table 1.**
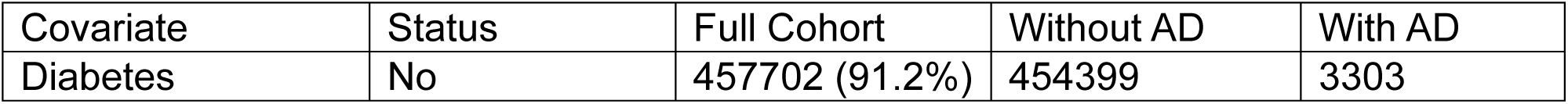
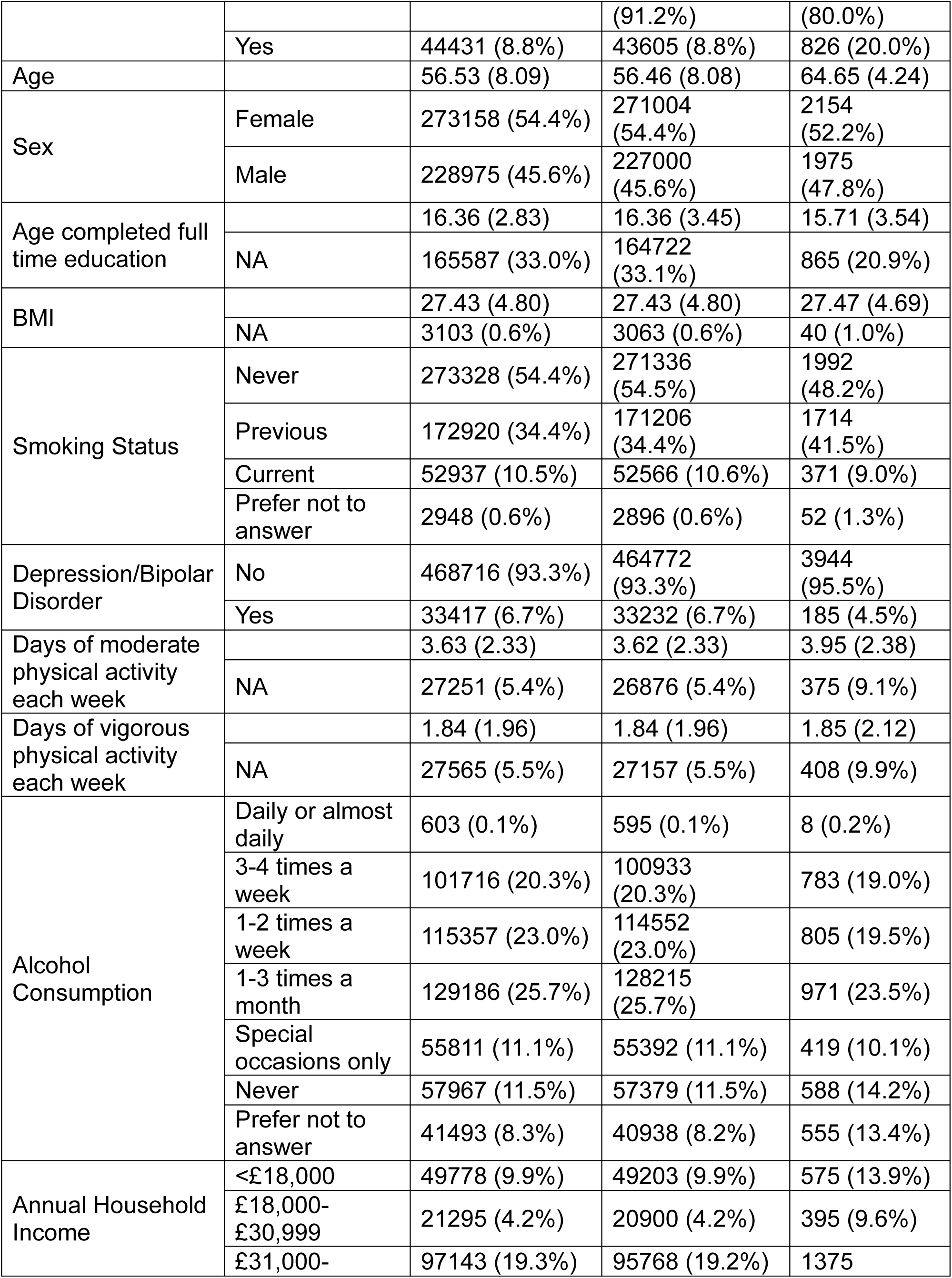
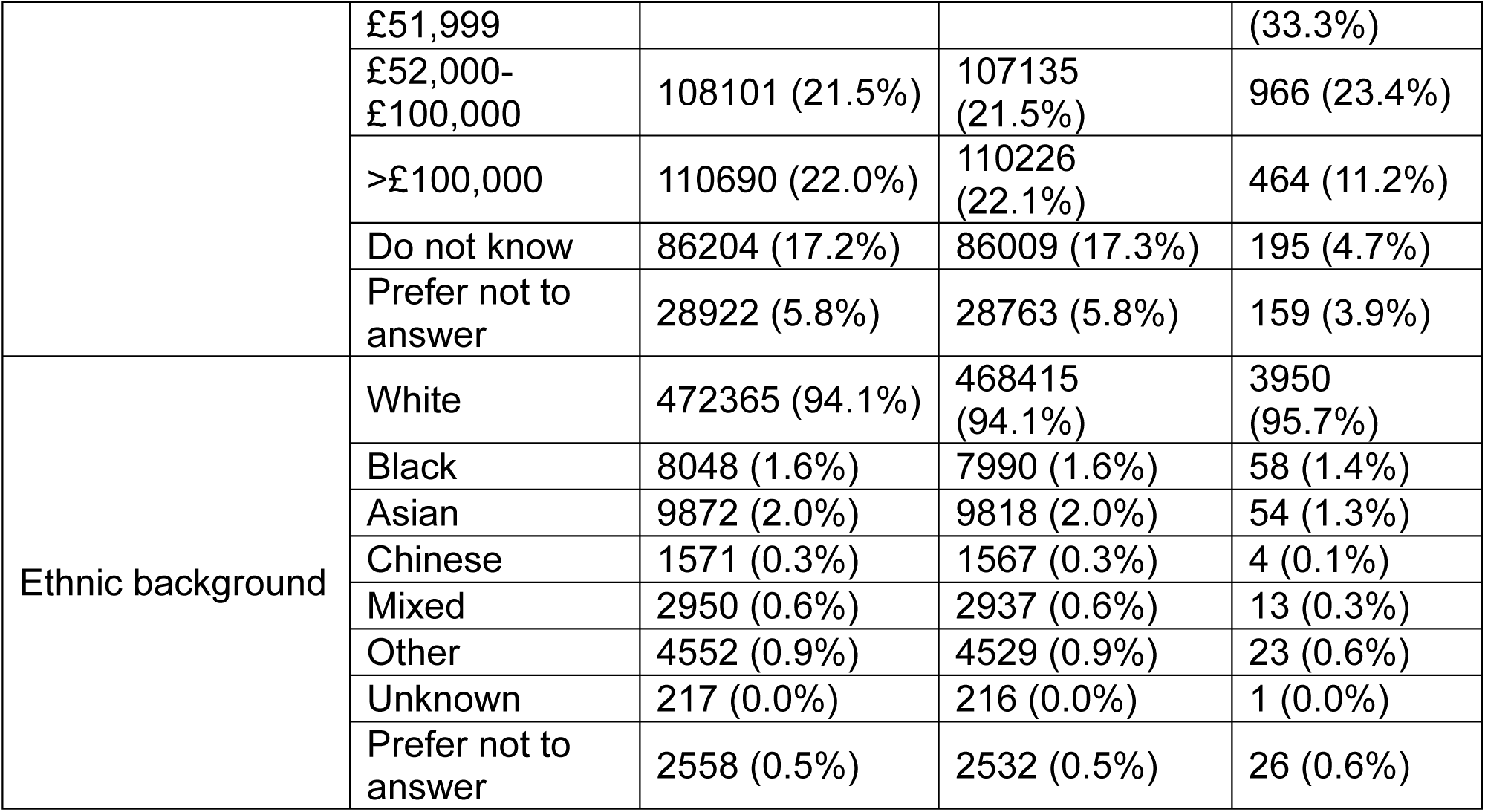
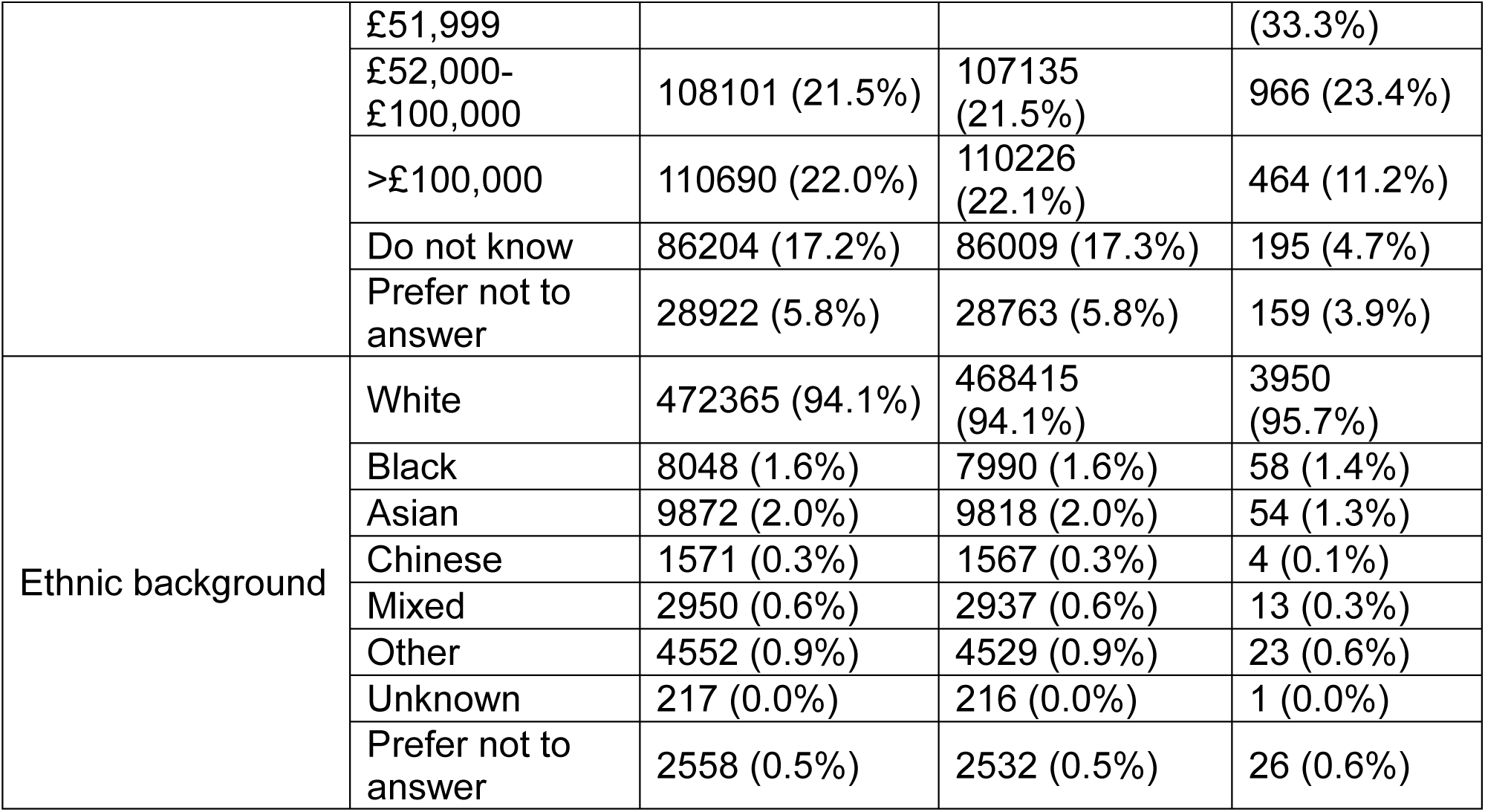
Demographic information about UKB full cohort and split by AD status.

**Table 2.** Demographic information about All of Us full cohort and split by AD status.

| Covariate | Status | Full Cohort | Without AD | With AD |
| --- | --- | --- | --- | --- |
| Diabetes | No | 237281 (82.7%) | 236807 (82.7%) | 474 (61.2%) |
|  | Yes | 49730 (17.3%) | 49430 (17.3%) | 300 (38.8%) |
| Age |  | 58.01 (16.96) | 57.95 (16.93) | 79.95 (9.88) |
| Sex | Female | 172401 (60.1%) | 171999 (60.1%) | 402 (51.9%) |
|  | Male | 108739 (37.9%) | 108388 (37.9%) | 351 (45.3%) |
|  | Intersex | 57 (0.0%) | 57 (0.0%) | 0 (0.0%) |
|  | No matching concept | 2702 (0.9%) | 2692 (0.9%) | 10 (1.3%) |
|  | None | 96 (0.0%) | 96 (0.0%) | 0 (0.0%) |
|  | Prefer not to answer | 3016 (1.1%) | 3005 (1.0%) | 11 (1.4%) |
| Highest Level of Education | Never Attended | 421 (0.1%) | 416 (0.1%) | 5 (0.6%) |
|  | Grades 1-4 | 2548 (0.9%) | 2532 (0.9%) | 16 (2.1%) |
|  | Grades 5-8 | 6745 (2.4%) | 6700 (2.3%) | 45 (5.8%) |
|  | Grades 9-11 | 18153 (6.3%) | 18115 (6.3%) | 38 (4.9%) |
|  | Grade 12 Or GED | 56778 (19.8%) | 56647 (19.8%) | 131 (16.9%) |
|  | 1-3 Years College | 72938 (25.4%) | 72780 (25.4%) | 158 (20.4%) |
|  | College Graduate | 62550 (21.8%) | 62394 (21.8%) | 156 (20.2%) |
|  | Advanced Degree | 57696 (20.1%) | 57494 (20.1%) | 202 (26.1%) |
|  | Prefer not to answer | 9182 (3.2%) | 9159 (3.2%) | 23 (3.0%) |
| BMI |  | 29.90 (7.64) | 29.90 (7.64) | 28.65 (6.57) |
|  | NA | 12715 (4.4%) | 12676 (4.4%) | 39 (5.0%) |
| Smoked 100 Cigarettes in Lifetime | No | 164021 (57.1%) | 163612 (57.2%) | 409 (52.8%) |
|  | Yes | 114595 (39.9%) | 114253 (39.9%) | 342 (44.2%) |
|  | Don't Know | 3645 (1.3%) | 3636 (1.3%) | 9 (1.2%) |
|  | Prefer not to answer | 4750 (1.7%) | 4736 (1.7%) | 14 (1.8%) |
| Depression | No | 208532 (72.7%) | 208193 (72.7%) | 339 (43.8%) |
|  | Yes | 78479 (27.3%) | 78044 (27.3%) | 435 (56.2%) |
| Frequency of Alcoholic Drinks over Past Year | 4 or More Per Week | 30087 (10.5%) | 30003 (10.5%) | 84 (10.9%) |
|  | 2 to 3 Per Week | 34981 (12.2%) | 34920 (12.2%) | 61 (7.9%) |
|  | 2 to 4 Per Month | 52212 (18.2%) | 52117 (18.2%) | 95 (12.3%) |
|  | Monthly Or Less | 84407 (29.4%) | 84226 (29.4%) | 181 (23.4%) |
|  | Never | 46653 (16.3%) | 46422 (16.2%) | 231 (29.8%) |
|  | Prefer not to answer | 38671 (13.5%) | 38549 (13.5%) | 122 (15.8%) |
| Annual Household Income | <\$10,000 | 40856 (14.2%) | 40781 (14.2%) | 75 (9.7%) |
| | \$10,000-\$25,000 | 33965 (11.8%) | 33851 (11.8%) | 114 (14.7%) |
| | \$25,000-\$35,000 | 20267 (7.1%) | 20216 (7.1%) | 51 (6.6%) |
| | \$35,000-\$50,000 | 22252 (7.8%) | 22184 (7.8%) | 68 (8.8%) |
| | \$50,000-\$75,000 | 29186 (10.2%) | 29106 (10.2%) | 80 (10.3%) |
| | \$75,000-\$100,000 | 22359 (7.8%) | 22308 (7.8%) | 51 (6.6%) |
| | \$100,000-\$150,000 | 27202 (9.5%) | 27146 (9.5%) | 56 (7.2%) |
| | \$150,000-\$200,000 | 12496 (4.4%) | 12472 (4.4%) | 24 (3.1%) |
| | >\$200,000 | 17507 (6.1%) | 17480 (6.1%) | 27 (3.5%) |
|  | Prefer not to answer | 60921 (21.2%) | 60693 (21.2%) | 228 (29.5%) |
| Ethnicity | White | 150402 (52.4%) | 149897 (52.4%) | 505 (65.2%) |
|  | Black | 57177 (19.9%) | 57088 (19.9%) | 89 (11.5%) |
|  | Hispanic | 47699 (16.6%) | 47579 (16.6%) | 120 (15.5%) |
|  | Asian | 8038 (2.8%) | 8025 (2.8%) | 13 (1.7%) |
|  | MENA | 1605 (0.6%) | 1603 (0.6%) | 2 (0.3%) |
|  | NHPI | 297 (0.1%) | 296 (0.1%) | 1 (0.1%) |
|  | Mixed | 10771 (3.8%) | 10755 (3.8%) | 16 (2.1%) |
|  | None Of These | 3017 (1.1%) | 3017 (1.1%) | 0 (0.0%) |
|  | Prefer not to answer | 8005 (2.8%) | 7977 (2.8%) | 28 (3.6%) |

### Cardiovascular Disease Subtypes and AD Prevalence

CVD subtypes showed varying prevalence across cohorts. In UKB, hypertension was most common (32.3%), followed by chronic ischemic heart disease and atrial fibrillation. In AoU, the distribution was similar, with slightly higher prevalence of hypertension (36.6%). AD prevalence was higher among individuals with each CVD subtype compared to those without, with the strongest differences seen in hypotension, heart failure, and cerebral infarction (Table 3 and Table 4).

**Table 3.** Counts of UKB participants with each subtype of CVD overall and split by AD status.

| CVD Subtype | CVD Status | n (%) | Without AD | With AD |
| --- | --- | --- | --- | --- |
| Hypertension | No | 339872 (67.7%) | 338205 (99.5%) | 1667 (0.5%) |
|  | Yes | 162261 (32.3%) | 159799 (98.5%) | 2462 (1.5%) |
| Hypotension | No | 480823 (95.8%) | 477440 (99.3%) | 3383 (0.7%) |
|  | Yes | 21310 (4.2%) | 20564 (96.5%) | 746 (3.5%) |
| Angina Pectoris | No | 467966 (93.2%) | 464473 (99.3%) | 3493 (0.7%) |
|  | Yes | 34167 (6.8%) | 33531 (98.1%) | 636 (1.9%) |
| Acute Myocardial Infarction | No | 484311 (96.5%) | 480433 (99.2%) | 3878 (0.8%) |
|  | Yes | 17822 (3.5%) | 17571 (98.6%) | 251 (1.4%) |
| Pulmonary Embolism | No | 492066 (98.0%) | 488111 (99.2%) | 3955 (0.8%) |
|  | Yes | 10067 (2.0%) | 9893 (98.3%) | 174 (1.7%) |
| Atrial Fibrillation | No | 460772 (91.8%) | 457474 (99.3%) | 3298 (0.7%) |
|  | Yes | 41361 (8.2%) | 40530 (98%) | 831 (2%) |
| Heart Failure | No | 482058 (96.0%) | 478397 (99.2%) | 3661 (0.8%) |
|  | Yes | 20075 (4.0%) | 19607 (97.7%) | 468 (2.3%) |
| Blockage | No | 488114 (97.2%) | 484345 (99.2%) | 3769 (0.8%) |
|  | Yes | 14019 (2.8%) | 13659 (97.4%) | 360 (2.6%) |
| Chronic Rheumatic Heart Disease | No | 490849 (97.8%) | 486930 (99.2%) | 3919 (0.8%) |
|  | Yes | 11284 (2.2%) | 11074 (98.1%) | 210 (1.9%) |
| Chronic Ischemic Heart Disease | No | 448703 (89.4%) | 445508 (99.3%) | 3195 (0.7%) |
|  | Yes | 53430 (10.6%) | 52496 (98.3%) | 934 (1.7%) |
| Cerebral Infarction | No | 491788 (97.9%) | 487900 (99.2%) | 3888 (0.8%) |
|  | Yes | 10345 (2.1%) | 10104 (97.7%) | 241 (2.3%) |

**Table 4.**
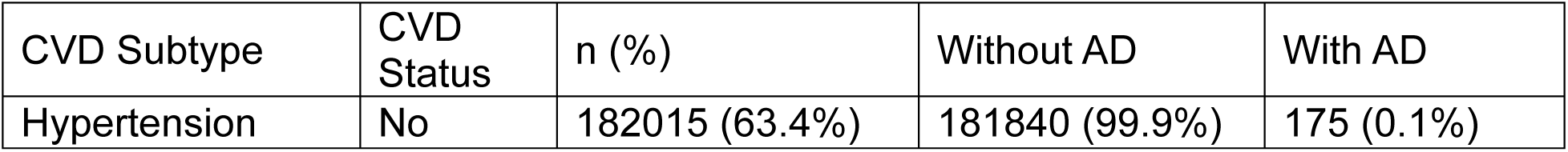
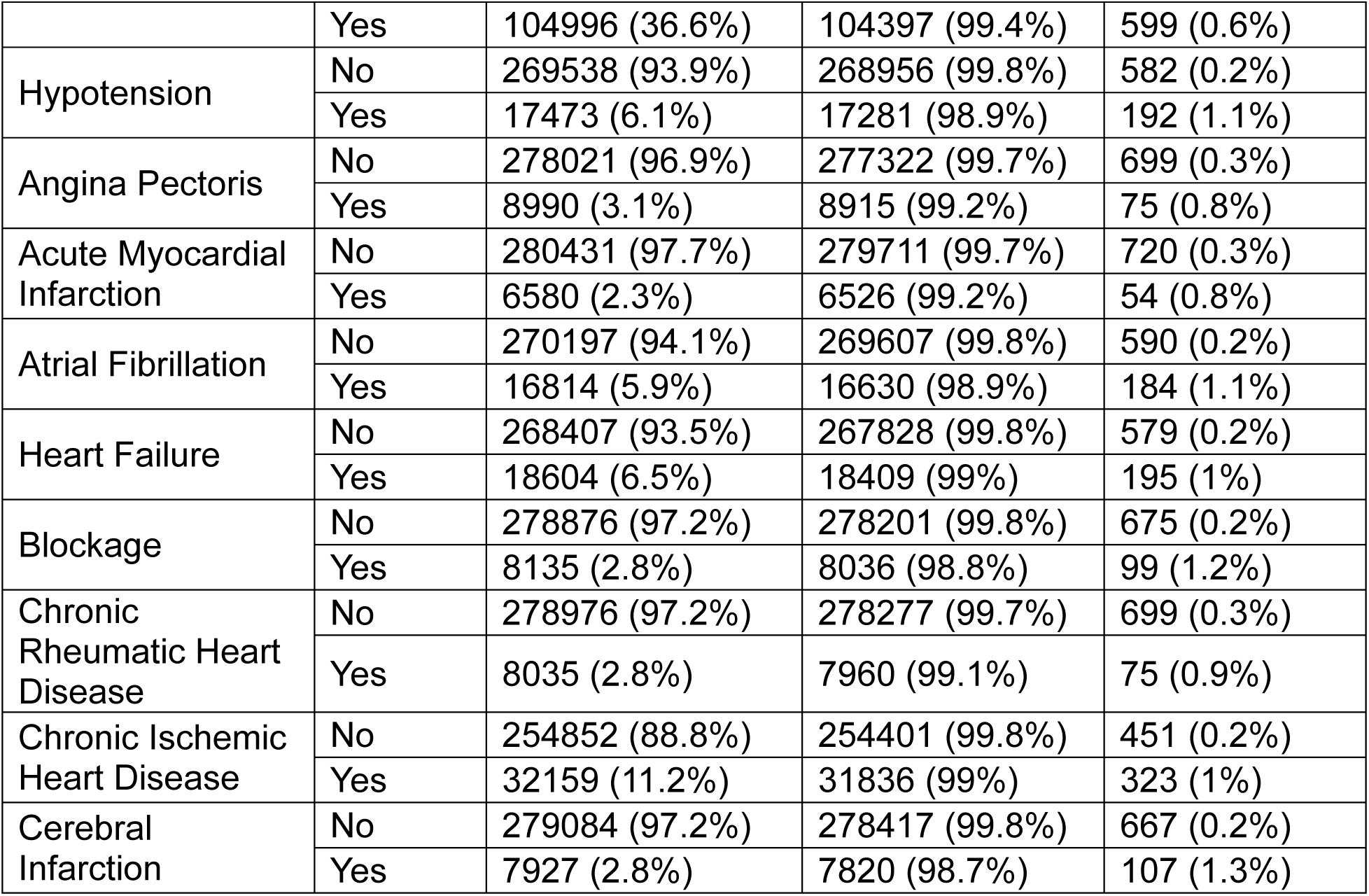
Counts of AoU participants with each subtype of CVD overall and split by AD status.

| CVD Subtype | CVD Status | n (%) | Without AD | With AD |
| --- | --- | --- | --- | --- |
| Hypertension | No | 182015 (63.4%) | 181840 (99.9%) | 175 (0.1%) |
|  | Yes | 104996 (36.6%) | 104397 (99.4%) | 599 (0.6%) |
| Hypotension | No | 269538 (93.9%) | 268956 (99.8%) | 582 (0.2%) |
|  | Yes | 17473 (6.1%) | 17281 (98.9%) | 192 (1.1%) |
| Angina Pectoris | No | 278021 (96.9%) | 277322 (99.7%) | 699 (0.3%) |
|  | Yes | 8990 (3.1%) | 8915 (99.2%) | 75 (0.8%) |
| Acute Myocardial Infarction | No | 280431 (97.7%) | 279711 (99.7%) | 720 (0.3%) |
|  | Yes | 6580 (2.3%) | 6526 (99.2%) | 54 (0.8%) |
| Atrial Fibrillation | No | 270197 (94.1%) | 269607 (99.8%) | 590 (0.2%) |
|  | Yes | 16814 (5.9%) | 16630 (98.9%) | 184 (1.1%) |
| Heart Failure | No | 268407 (93.5%) | 267828 (99.8%) | 579 (0.2%) |
|  | Yes | 18604 (6.5%) | 18409 (99%) | 195 (1%) |
| Blockage | No | 278876 (97.2%) | 278201 (99.8%) | 675 (0.2%) |
|  | Yes | 8135 (2.8%) | 8036 (98.8%) | 99 (1.2%) |
| Chronic Rheumatic Heart Disease | No | 278976 (97.2%) | 278277 (99.7%) | 699 (0.3%) |
|  | Yes | 8035 (2.8%) | 7960 (99.1%) | 75 (0.9%) |
| Chronic Ischemic Heart Disease | No | 254852 (88.8%) | 254401 (99.8%) | 451 (0.2%) |
|  | Yes | 32159 (11.2%) | 31836 (99%) | 323 (1%) |
| Cerebral Infarction | No | 279084 (97.2%) | 278417 (99.8%) | 667 (0.2%) |
|  | Yes | 7927 (2.8%) | 7820 (98.7%) | 107 (1.3%) |

**Table 5.**
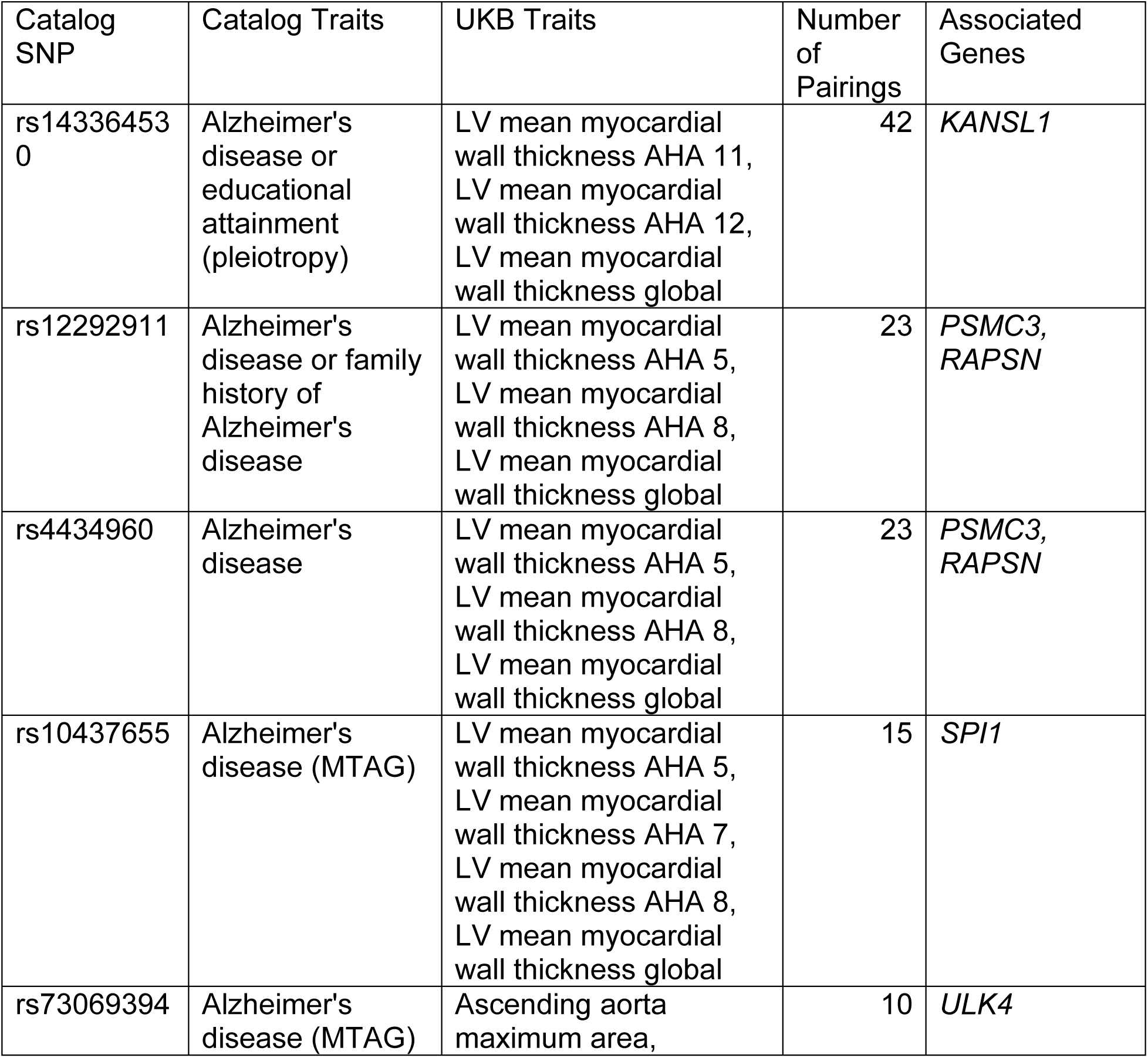
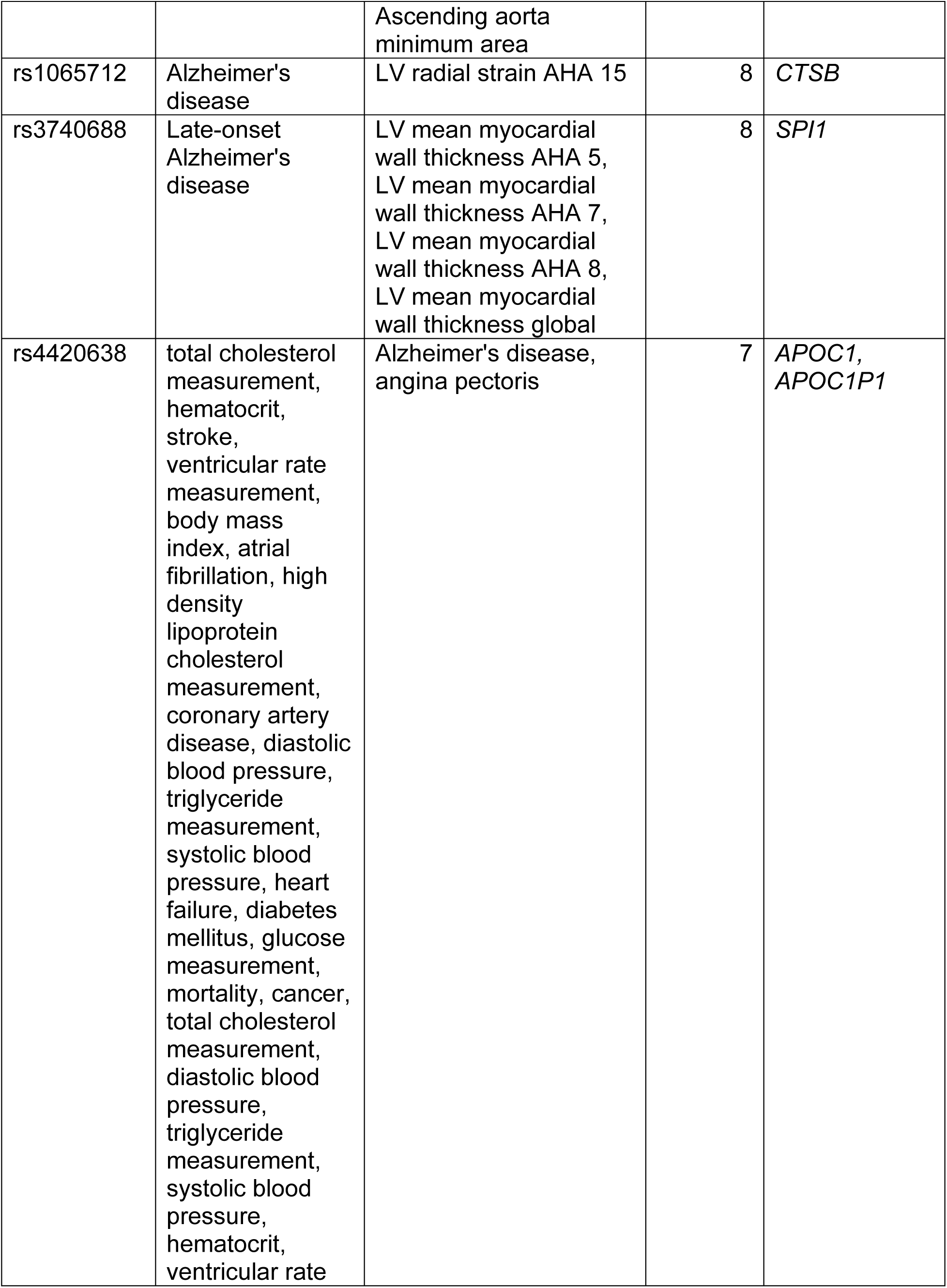
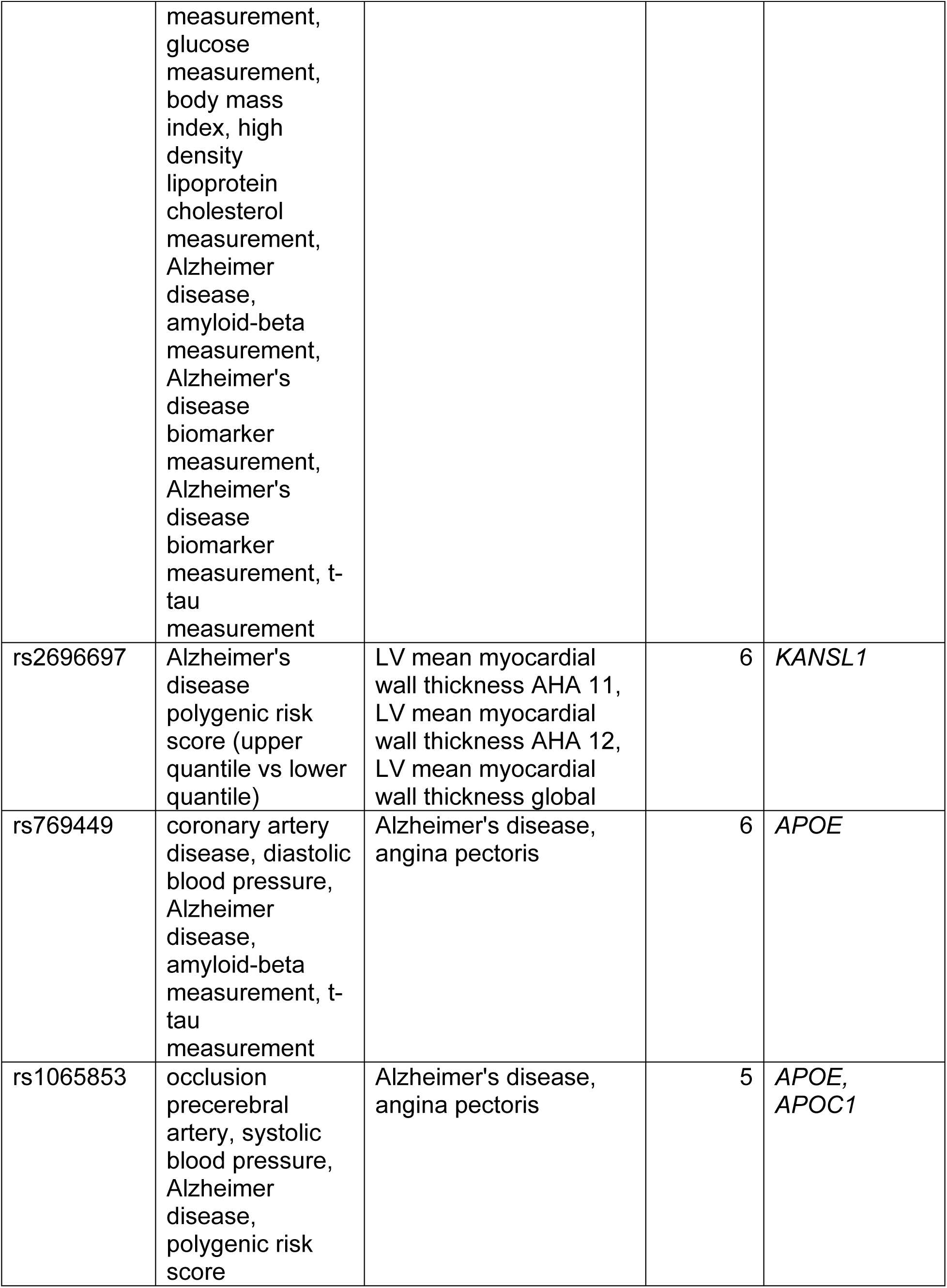
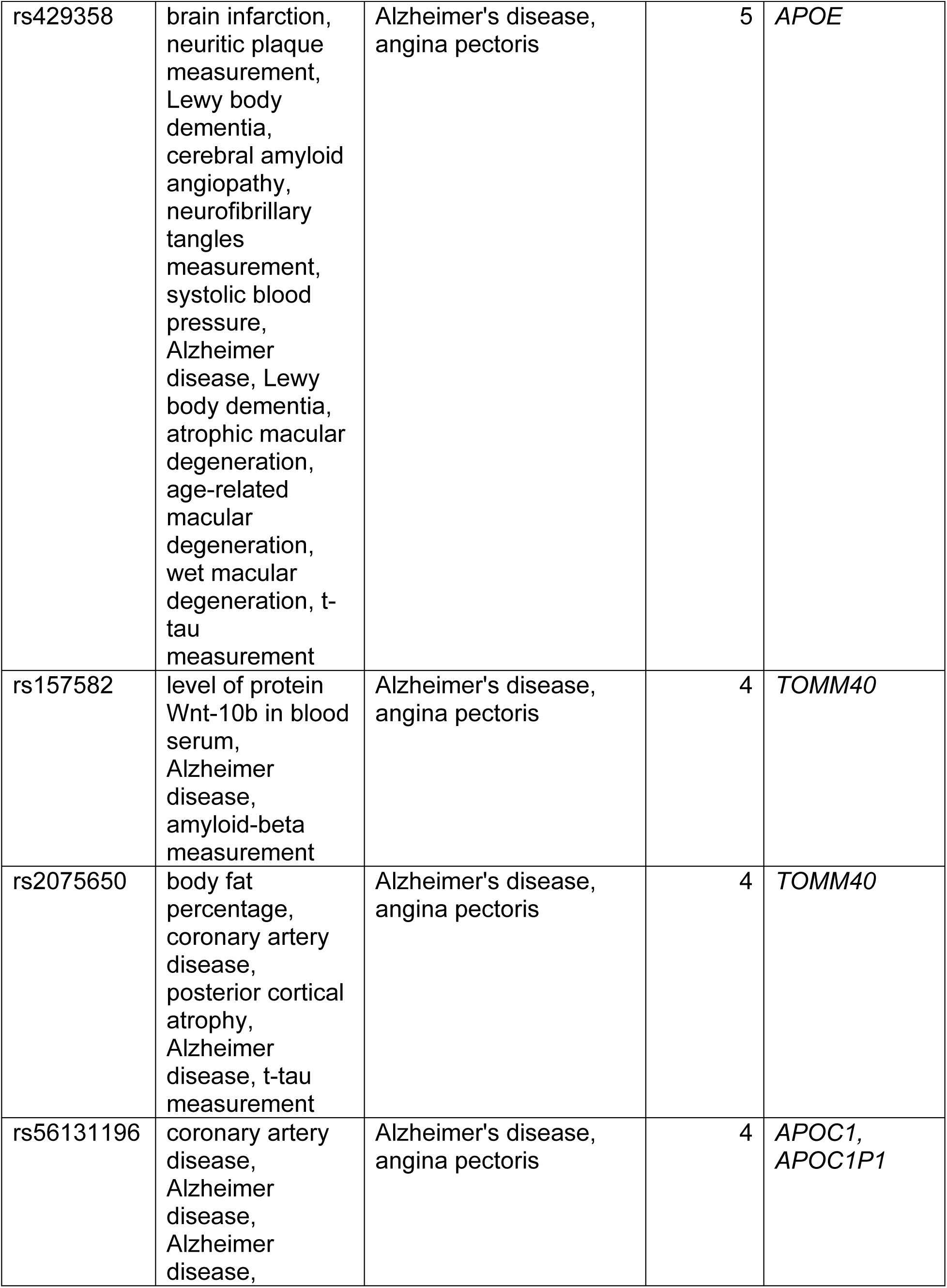
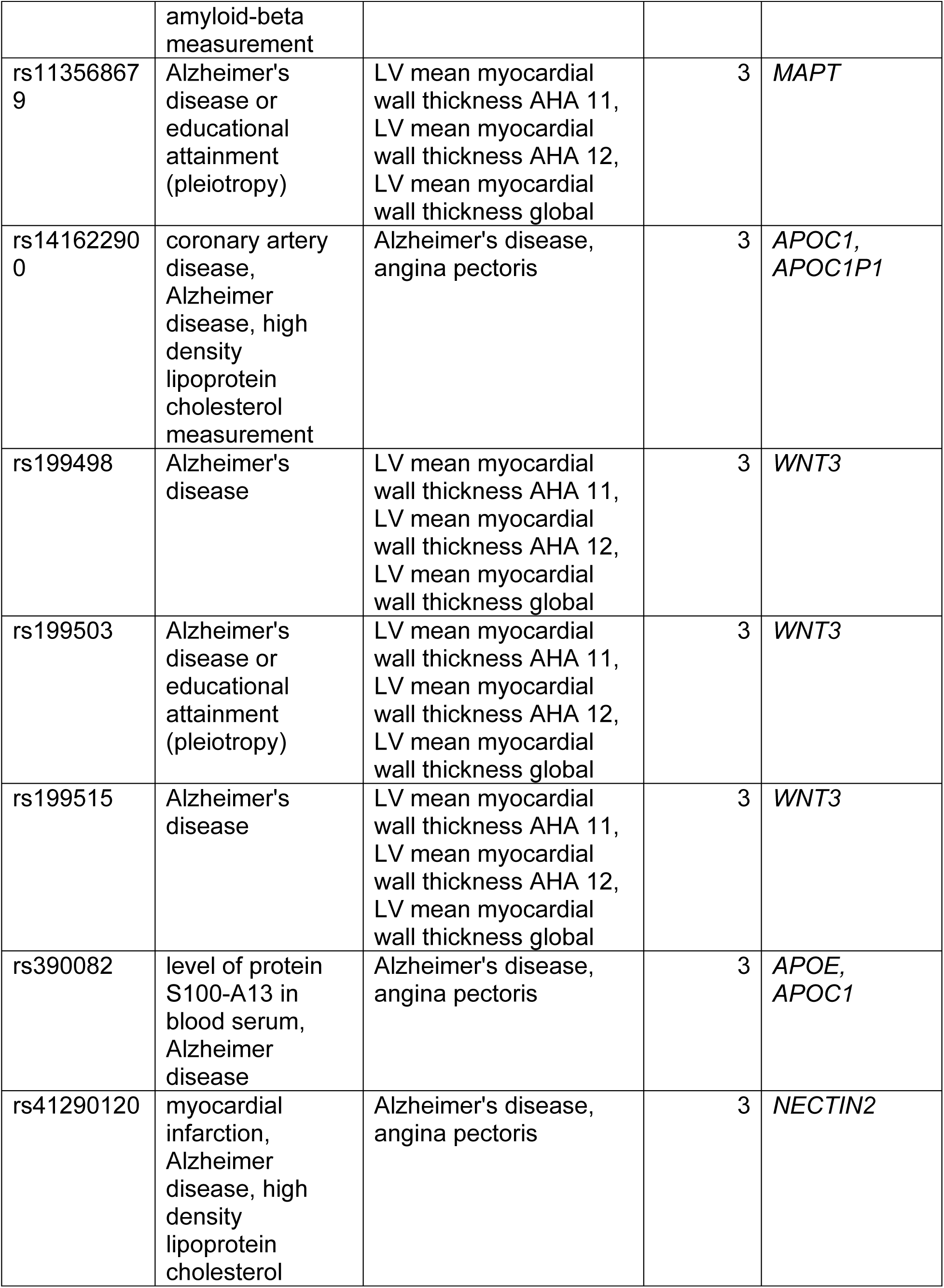
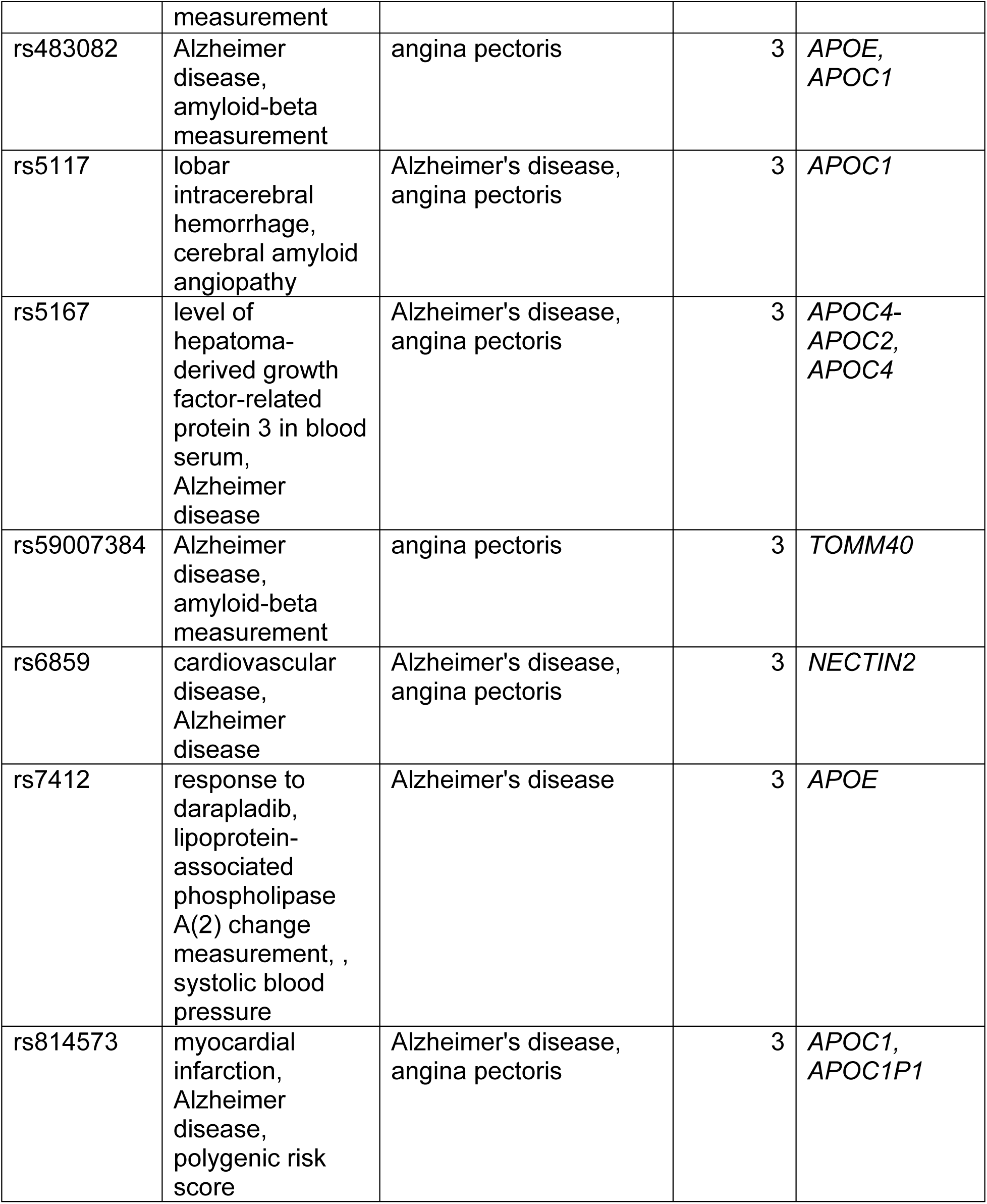
Proximal SNPs related to the heart and brain. Traits are reported associated with SNP in the catalog, as well as traits that had significant UKB GWAS results, the number of pairings with each catalog SNP, and the gene(s) associated with each SNP. Only SNPs with greater than 2 pairings are reported.

### Associations Between CVD Subtypes and AD

Logistic regression analysis adjusted for relevant covariates revealed that nearly all CVD subtypes were significantly associated with higher odds of AD in both cohorts (Figure 1 and Figure 2). In UKB, the strongest association was observed between hypotension and AD (OR = 2.74, 95% CI: [2.52, 2.98]), followed by blockages (OR = 1.62, 95% CI: [1.45, 1.82]), hypertension (OR = 1.57, 95% CI: [1.47, 1.68]), and cerebral infarction (OR = 1.49, 95% CI: [1.30, 1.71]). Acute myocardial infarction was the only subtype not significantly associated with AD (OR = 1.01, 95% CI: [0.89, 1.15]).

**Figure 1.**
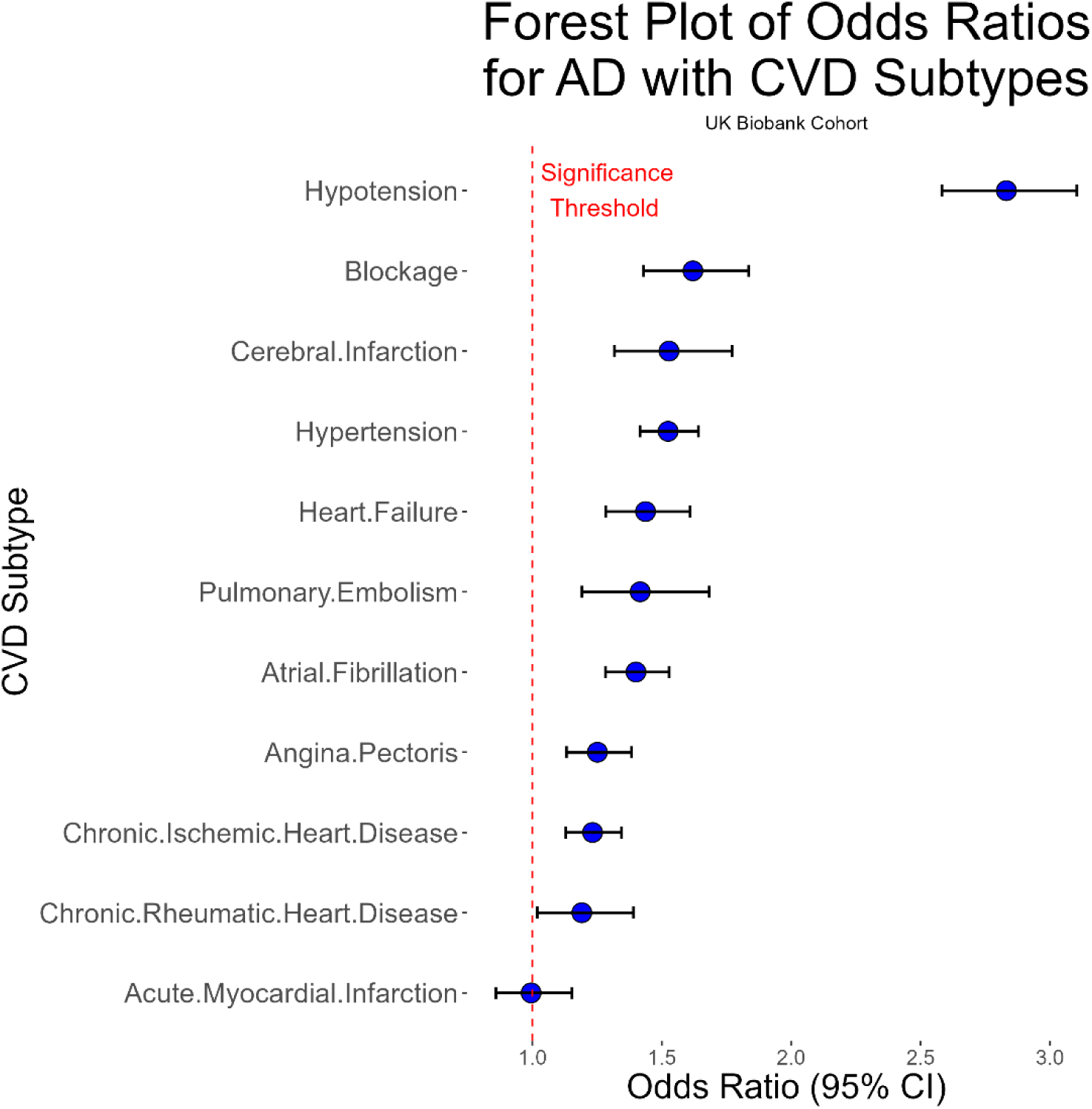
Forest plot of odds ratios for AD with CVD subtypes in the UKB cohort. Blue dots represent odds ratio point estimates, and error bars show the 95% confidence interval. The dotted red line shows the significance threshold. By far the strongest association was noted between hypotension and AD. All CVD subtypes besides acute myocardial infarction had significant relationships with AD.

**Figure 2.**
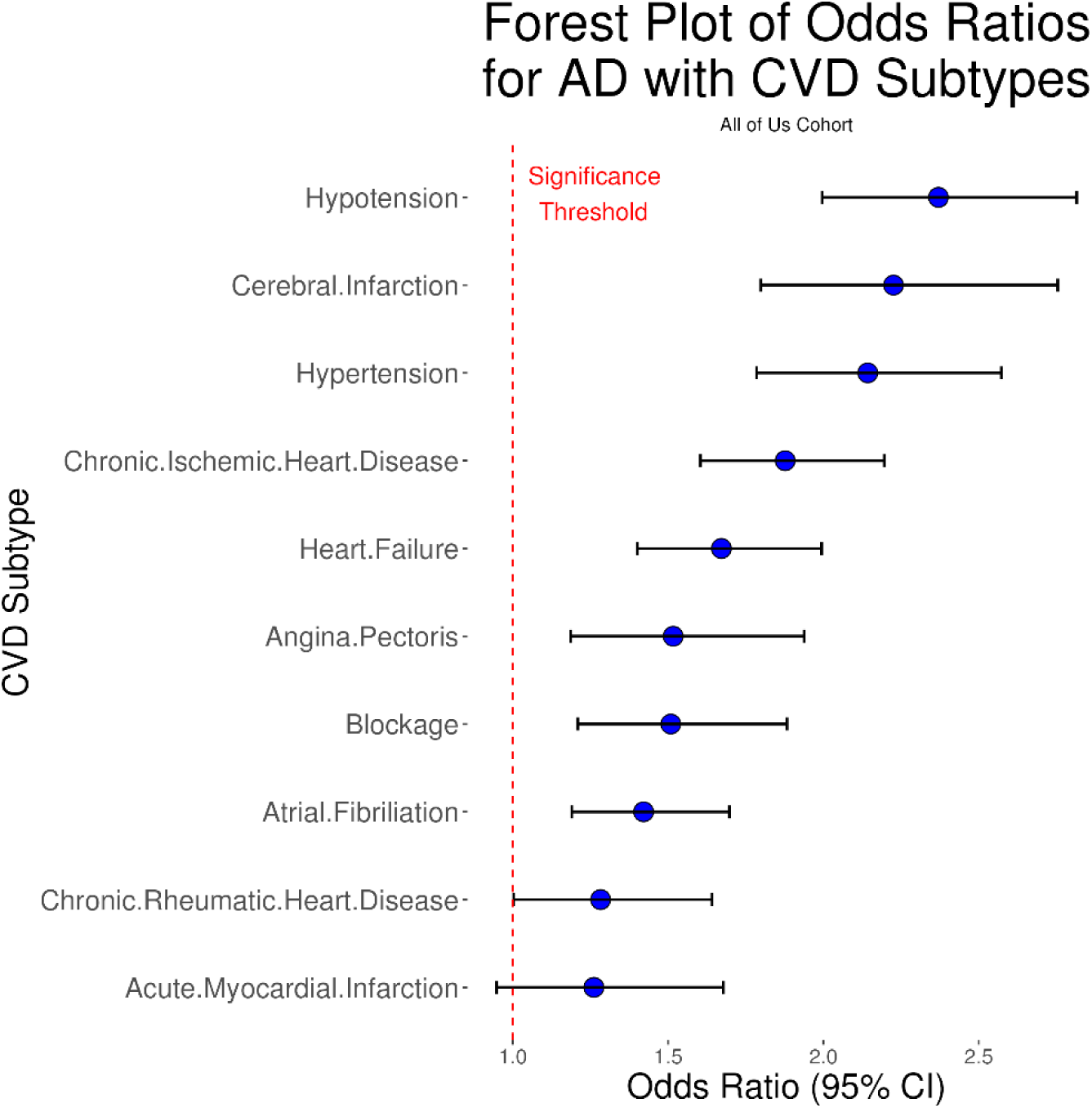
Forest plot of odds ratios for AD with CVD subtypes in the All of Us cohort. Blue dots represent odds ratio point estimates, and error bars show the 95% confidence interval. The dotted red line shows the significance threshold. Note the strongest associations between AD and hypotention, cerebral infarction, and hypertension. All CVD subtypes were significant at the 95% confidence level except for acute myocardial infarction.

Findings in AoU mirrored those from UKB. Again, hypotension showed the strongest association with AD (OR = 2.37, 95% CI: [2.00, 2.82], followed by cerebral infarction (OR = 2.23, 95% CI: [1.80, 2.75]) and hypertension (OR = 2.14, 95% CI: [1.78, 2.57]). AMI remained non-significant (OR = 1.26, 95% CI: [0.95, 1.68]). Odds ratio estimates in AoU had wider confidence intervals, likely reflecting the greater racial/ethnic diversity, broader socioeconomic representation, and the lack of adjustment for physical activity.

### Genetic Overlap Between CVD and AD

To explore shared genetic architecture, we identified SNPs significantly associated with both CVD and AD-related traits that were located within 50 kb of one another (p < 5×10⁻⁸). A total of 164 unique SNP pairs were found to be proximal across datasets, with the strongest overlaps observed between AD and traits such as angina pectoris, left ventricular myocardial wall thickness, and coronary artery disease.

Several loci stood out for high overlap, including:

- **19q13.32**, encompassing *APOE*, *TOMM40*, *APOC1*, *APOC1P1*, *NECTIN2*, and *APOC4-APOC2*, linked to lipid metabolism and both AD and cardiovascular traits^27–30^.
- **17q21.31**, home to *MAPT*, *KANSL1*, and *WNT3* with links to ventricular wall thickness and AD risk^25^.
- **11p11.2**, containing *PSMC3*, *SPI1*, and *RAPSN*, genes implicated in neuroinflammation, immune response, and cardiac structure^31,32^.

## Discussion

This study used two large, demographically distinct biobank datasets—UKB and AoU—to investigate associations between AD and multiple CVD subtypes. Our findings demonstrate that most CVD subtypes are significantly associated with higher odds of AD, with hypotension emerging as the strongest and most consistent correlation in both cohorts. These results highlight the importance of examining the heart–brain axis beyond general CVD risk and underscore the value of differentiating CVD subtypes in dementia research.

Notably, hypotension showed the highest odds of AD across both datasets. While hypertension has been extensively studied as a modifiable AD risk factor, hypotension is comparatively understudied despite its high prevalence among older adults^33^. Both chronic and orthostatic hypotension have been associated with cerebral hypoperfusion, oxidative stress, and tau pathology—mechanisms that could exacerbate or accelerate AD progression^2,13^. Conversely, AD-related dysfunction of the autonomic nervous system may impair cardiovascular regulation, suggesting a bidirectional relationship^34^.

Hypertension and cerebral infarction also demonstrated strong associations with AD, consistent with established literature linking these conditions to vascular pathology^2,4,15,35,36^, white matter damage^7^, and cognitive decline^5,37^. Atrial fibrillation, known to increase stroke risk and independently linked to cognitive impairment, was similarly associated with AD^16,38^. These findings support a broader vascular contribution to AD and indicate that multiple CVD pathways, particularly those impacting cerebral blood flow, may converge in AD pathophysiology.

The only CVD subtype not significantly associated with AD was acute myocardial infarction (AMI). This aligns with prior research suggesting that while AMI may contribute to cognitive decline through systemic inflammation or hypoxia^39^, its direct link to AD pathology remains limited^14,40^. However, AMI’s indirect effects on brain health warrant further investigation, particularly regarding rates of long-term cognitive decline post-infarction^39,41,42^.

Our genetic colocalization analysis identified overlapping SNPs between CVD and AD traits, especially in loci involving *APOE*, *MAPT*, *SPI1*, and *WNT3*. The *APOE* region (19q13.32) remains the most prominent shared locus, given its well-established roles in lipid metabolism^27,29^, blood-brain barrier integrity^6,28^, and both cardiovascular and neurodegenerative diseases^27,28,30^. Interestingly, we also observed overlap between myocardial wall thickness traits and AD-related SNPs in several regions, including 17q21.31 (*MAPT*) and 11p11.2 (*PSMC3*, *RAPSN*), pointing to potential shared mechanisms involving cardiac remodeling and brain structure integrity^25,43,44^.

Differences between the two cohorts—UKB’s healthier, less diverse sample versus AoU’s more representative and clinically diverse population—underscore the generalizability of our findings. The replicated associations across these distinct cohorts strengthen the robustness of our results and highlight the need for tailored prevention strategies that address specific CVD subtypes in diverse populations.

Despite the strengths of large-scale, multi-cohort replication and integration of genetic data, this study has limitations. First, its cross-sectional design precludes causal inference, and the directionality of associations cannot be determined. Second, AD and CVD diagnoses were based on ICD-10 codes, which may underrepresent true disease prevalence due to undiagnosed or misclassified cases. Third, we did not adjust for cardiovascular multimorbidity, and participants with multiple CVD subtypes may have higher cumulative AD risk^45,46^. Additionally, our genetic analysis, while informative, used proximity-based colocalization rather than formal methods such as Mendelian randomization or transcriptome-wide association studies, which could better infer causality or gene expression effects.

## Conclusion

In this study, we investigated the relationship between AD and multiple CVD subtypes across two large, diverse biobank cohorts: the UK Biobank and All of Us. We found that most CVD subtypes were significantly associated with AD, with hypotension showing the strongest association in both datasets. Other conditions like hypertension, cerebral infarction, and atrial fibrillation also demonstrated consistent positive associations, reinforcing the central role of vascular health in cognitive decline. Acute myocardial infarction was the only subtype not significantly linked to AD, consistent with prior literature.

Genetic analyses revealed that several AD- and CVD-associated SNPs were located near each other, especially in regions containing *APOE*, *MAPT*, and genes involved in myocardial structure. These results suggest a potential shared genetic basis between heart and brain pathology. While the cross-sectional design and reliance on ICD-10 codes limit causal inference, our findings underscore the importance of cardiovascular health in AD risk and highlight specific CVD subtypes that may warrant increased attention in prevention strategies.

## Data Availability

This research has been conducted using the UK Biobank Resource under Application Number 61915.We also thank the National Institutes of Health?s All of Us Research Program for making available the participant data examined in this study. The computer source code for this study is available at https://github.com/MIILab-MTU/CVDSubtypesADCorrelation.

## Data Availability Statement

The computer source code for this study is available at https://github.com/MIILab-MTU/CVDSubtypesADCorrelation.

## Acknowledgements

This research has been conducted using the UK Biobank Resource under Application Number 61915. We gratefully acknowledge the UK Biobank and All of Us participants for their contributions, without whom this research would not have been possible. We also thank the National Institutes of Health’s All of Us Research Program for making available the participant data examined in this study.

